# Quantifying the health impact of food interventions: Revisiting the Disability-Adjusted Life Years Approach

**DOI:** 10.1101/2024.08.26.24312574

**Authors:** Bert Lenaerts

## Abstract

Food interventions like industrial fortification and biofortification through crop breeding can help shift towards improved healthy diets, marking a significant stride in public health. Crop breeding contributes to a stable and healthy food supply by boosting agricultural yields and the micronutrient content of staples, which is pivotal for combating chronic and hidden hunger, especially in rural areas. Fortification enhances healthy diets by adding essential vitamins and minerals to commonly consumed foods, helping prevent nutrient deficiencies and support overall well-being. The burden of hunger and its consequences on health are increasingly quantified using the Disability-Adjusted Life Years (DALYs) approach, which merges years of life lost and years lived with disability, offering a comprehensive view of health impacts and aiding in resource allocation. A practical formula for quantifying the health impact of biofortification was introduced by Stein et al. (2005) and Zimmermann and Qaim (2004). This entails calculating the efficacy or relative reduction in hunger burden based on the current and post-intervention nutrient intake against the recommended dietary allowances. As data on consumption and recommended intake levels are variable and not robustly available, this paper proposes relying on relative estimates to bridge the data gaps and uncertainties, thus streamlining the quantification of fortification’s and biofortification’s impact on diets and overall health.

## 1 Classic DALY approach

Crop breeding increases yields, enhances disease resistance, and improves climate resilience, ensuring a stable food supply. Besides boosting yields, crop breeding can increase the micronutrient content of staple crops through biofortification (Bouis et al., 2011). Fortification works by incorporating targeted nutrients like iron, iodine, folic acid, or vitamin D into staple foods—such as flour, salt, milk, or cereals—during manufacturing, ensuring widespread access to essential micronutrients. These commodity improvements can help combat chronic and hidden hunger and provide a sustainable way to improve nutrition through regular diets, especially in rural areas.

The burden of hunger is increasingly being measured in Disability-Adjusted Life Years (DALYs). DALYs measure disease burden by combining years of life lost (YLL) and years lived with disability (YLD). They provide a comprehensive view of health impacts, enabling comparison across diseases and aiding resource allocation. However, DALYs are complex to calculate, involve subjective disability weights, and focus on adverse health aspects. Despite these drawbacks, DALYs are valuable for understanding and addressing public health priorities.

Stein et al. (2005) and Zimmermann and Qaim (2004) pioneered a practical formula for quantifying the health impact of biofortification:

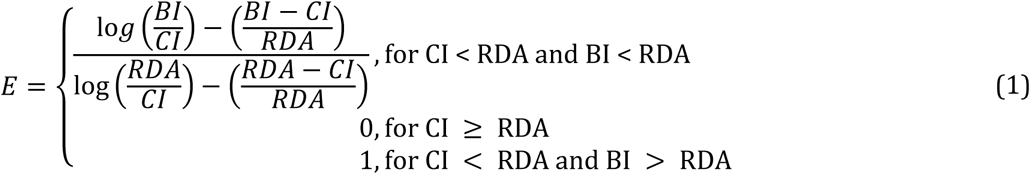

where E is the efficacy or relative reduction in the burden of hunger, CI is the current intake, BI is the intake after intervention (such as biofortification), and RDA is the recommended dietary allowance for a specific nutrient (see Table S1 for an index of terms). Although Stein et al. (2005) and Zimmermann and Qaim (2004) applied their method to the biofortification of micronutrients (iron, zinc, vitamin A), equation 1 also applies to macronutrients (calories and proteins).

The efficacy rate can then be used to quantify the (absolute) reduction in the burden of hunger (compared to the baseline or Business As Usual (BUA) scenario):

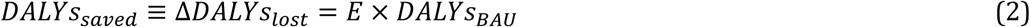

## 2 Modified DALY approach

Operationalising Equation 2 requires estimates of CI, BI, RDA and DALYs. The Institute for Health Metrics and Evaluation provides the health burden (expressed in DALYs) of different nutrient deficiencies. Estimates of CI and RDA are widely reported but exhibit considerable variation across sources (Lividini and Masters, 2022; Passarelli et al., 2024; Safiri et al., 2021; Wang et al., 2023; Wessells and Brown, 2012). Quantifying CI requires commodity-specific accounting for (1) the nutrient supply through production, trade, waste and non-food usage, (2) nutrient-specific losses through processing and cooking and (3) bioavailability through chemical composition, food interactions, and human absorption capacity. RDA captures the nutrient intake needed to meet the requirements of healthy individuals and can be obtained from different agencies (US National Institutes of Health,

European Food Safety Authority, UN Food and Agriculture Organization). However, different versions exist (minimum dietary requirement, average dietary requirement and adequate intake), reflecting the distribution across demographies, complicating comparisons across sources.

We propose to use relative estimates of CI, BI and RDA to overcome existing data gaps and uncertainty:

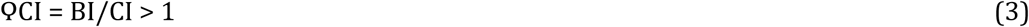

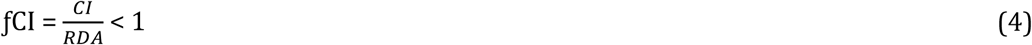

where ϘCI is the relative nutrient intake increase and ƒCI is the relative nutrient intake gap (or fraction of recommended intake that is consumed).

Using relative metrics helps overcome data gaps and increases robustness and accuracy. First, relative parameters maintain consistent measurements within a dataset even with systematic errors or uncertainties by reducing the impact of (absolute) outliers, leading to more robust results (see Section on Relative nutrient intake gaps). Second, relative parameters enable comparison across different nutrients (calories, proteins, iron, zinc, vitamin A). Third, relative parameters ensure that repeated measurements are consistent, which is crucial for modelling impact across multiple crops and geographies. Last, relative parameters can be adjusted and fine-tuned more easily without recalibrating the entire system.

### 2.1 Baseline health burden of hunger

The Institute for Health Metrics and Evaluation (IHME, 2024a) provides the health burden (expressed in DALYs) of different nutrient deficiencies (calories/proteins, iron, zinc, vitamin A, iodine) disaggregated by country, year, gender and age via the Global Burden of Disease (GBD) database (Table 1). Nutritional deficiencies are captured as cause and/or risk factors. Cause factors here are hunger-related diseases or injuries (that is, nutritional deficiencies) that directly cause death or disability; risk factors (per cause-risk attribution) are hunger-related factors that are causally associated with the probability of a disease or injury (Gödecke et al., 2018; Lenaerts and Demont, 2021). We recommend risk factors (if available) since they capture multiple cause factors, including indirect ones (like maternal death linked to iron deficiency) (see Eq. 9 and Table S1).

**Table 1.** Overview of global hunger-related risk and cause factors by age group for wasting, iron, vitamin A and zinc in 2021.

| Risk factor | Cause factor | Age group | DALY |
| --- | --- | --- | --- |
| Child wasting | Diarrheal diseases | 0-4 years | 1.59E7 |
| Child wasting | Lower respiratory infections | 0-4 years | 1.01E7 |
| Child wasting | Measles | 0-4 years | 1.07E6 |
| Child wasting | Protein-energy malnutrition | 0-4 years | 7.93E6 |
| Child wasting | Protein-energy malnutrition | 5-9 years | 431,719 |
| Child wasting | Protein-energy malnutrition | 10+ years | 3.45E6 |
| Iron deficiency | Dietary iron deficiency | 0-4 years | 5.73E6 |
| Iron deficiency | Dietary iron deficiency | 5-9 years | 4.12E6 |
| Iron deficiency | Dietary iron deficiency | 10+ years | 2.24E7 |
| Vitamin A deficiency | Diarrheal diseases | 0-4 years | 1.01E6 |
| Vitamin A deficiency | Measles | 0-4 years | 548,495 |
| Vitamin A deficiency | Vitamin A deficiency | 0-4 years | 478,299 |
| Vitamin A deficiency | Vitamin A deficiency | 5-9 years | 285,005 |
| Vitamin A deficiency | Vitamin A deficiency | 10+ years | 340,940 |
| Zinc deficiency | Diarrheal diseases | 0-4 years | 1.05E06 |
| Zinc deficiency | Other causes | 0-4 years | 1.05E06 |

Source: GBD 2021 from IHME (2024)

**Table 2.**
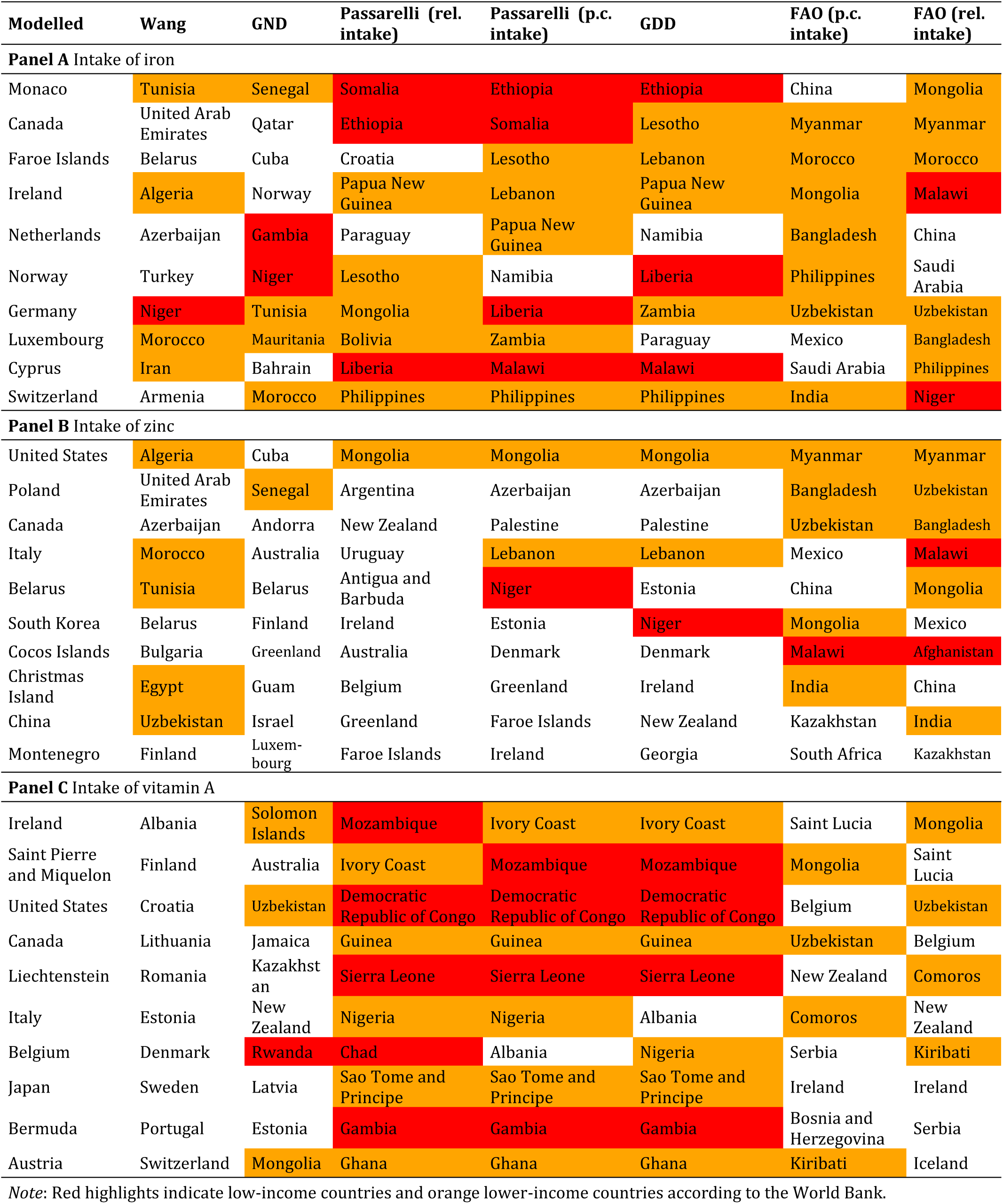
Top 10 countries with the highest intake of iron, zinc and vitamin A (beta-carotene).

| Modelled | Wang | GND | Passarelli (rel. intake) | Passarelli (p.c. intake) | GDD | FAO (p.c. intake) | FAO (rel. intake) |
| --- | --- | --- | --- | --- | --- | --- | --- |
| **Panel A** Intake of iron |  |  |  |  |  |  |  |
| Monaco | Tunisia | Senegal | Somalia | Ethiopia | Ethiopia | China | Mongolia |
| Canada | United Arab Emirates | Qatar | Ethiopia | Somalia | Lesotho | Myanmar | Myanmar |
| Faroe Islands | Belarus | Cuba | Croatia | Lesotho | Lebanon | Morocco | Morocco |
| Ireland | Algeria | Norway | Papua New Guinea | Lebanon | Papua New Guinea | Mongolia | Malawi |
| Netherlands | Azerbaijan | Gambia | Paraguay | Papua New Guinea | Namibia | Bangladesh | China |
| Norway | Turkey | Niger | Lesotho | Namibia | Liberia | Philippines | Saudi Arabia |
| Germany | Niger | Tunisia | Mongolia | Liberia | Zambia | Uzbekistan | Uzbekistan |
| Luxembourg | Morocco | Mauritania | Bolivia | Zambia | Paraguay | Mexico | Bangladesh |
| Cyprus | Iran | Bahrain | Liberia | Malawi | Malawi | Saudi Arabia | Philippines |
| Switzerland | Armenia | Morocco | Philippines | Philippines | Philippines | India | Niger |
| **Panel B** Intake of zinc |  |  |  |  |  |  |  |
| United States | Algeria | Cuba | Mongolia | Mongolia | Mongolia | Myanmar | Myanmar |
| Poland | United Arab Emirates | Senegal | Argentina | Azerbaijan | Azerbaijan | Bangladesh | Uzbekistan |
| Canada | Azerbaijan | Andorra | New Zealand | Palestine | Palestine | Uzbekistan | Bangladesh |
| Italy | Morocco | Australia | Uruguay | Lebanon | Lebanon | Mexico | Malawi |
| Belarus | Tunisia | Belarus | Antigua and Barbuda | Niger | Estonia | China | Mongolia |
| South Korea | Belarus | Finland | Ireland | Estonia | Niger | Mongolia | Mexico |
| Cocos Islands | Bulgaria | Greenland | Australia | Denmark | Denmark | Malawi | Afghanistan |
| Christmas Island | Egypt | Guam | Belgium | Greenland | Ireland | India | China |
| China | Uzbekistan | Israel | Greenland | Faroe Islands | New Zealand | Kazakhstan | India |
| Montenegro | Finland | Luxembourg | Faroe Islands | Ireland | Georgia | South Africa | Kazakhstan |
| **Panel C** Intake of vitamin A |  |  |  |  |  |  |  |
| Ireland | Albania | Solomon Islands | Mozambique | Ivory Coast | Ivory Coast | Saint Lucia | Mongolia |
| Saint Pierre and Miquelon | Finland | Australia | Ivory Coast | Mozambique | Mozambique | Mongolia | Saint Lucia |
| United States | Croatia | Uzbekistan | Democratic Republic of Congo | Democratic Republic of Congo | Democratic Republic of Congo | Belgium | Uzbekistan |
| Canada | Lithuania | Jamaica | Guinea | Guinea | Guinea | Uzbekistan | Belgium |
| Liechtenstein | Romania | Kazakhstan | Sierra Leone | Sierra Leone | Sierra Leone | New Zealand | Comoros |
| Italy | Estonia | New Zealand | Nigeria | Nigeria | Albania | Comoros | New Zealand |
| Belgium | Denmark | Rwanda | Chad | Albania | Nigeria | Serbia | Kiribati |
| Japan | Sweden | Latvia | Sao Tome and Principe | Sao Tome and Principe | Sao Tome and Principe | Ireland | Ireland |
| Bermuda | Portugal | Estonia | Gambia | Gambia | Gambia | Bosnia and Herzegovina | Serbia |
| Austria | Switzerland | Mongolia | Ghana | Ghana | Ghana | Kiribati | Iceland |

Note: Red highlights indicate low-income countries and orange lower-income countries according to the World Bank.

Following Caulfield et al. (2006), we consider wasting a proxy for macronutrient (calories and proteins) deficiency. Wasting results from inadequate nutrition over a shorter period, and is therefore impactable by increases in agricultural production. Stunting, resulting from longer-term undernutrition, is also driven by underlying causes like bad sanitation and extreme poverty, which are not directly solved by higher calorie or protein yields.

Following Gödecke et al. (2018), we combine iodine (ID), iron (Fe), zinc (Zn), vitamin A (VA) and other nutrient (Other) deficiencies (D) into hidden hunger. Unlike Gödecke et al. (2018), we exclude maternal diseases since these are not related to iron intake from food. We keep zinc deficiency (risk factor), although it is partially included in other deficiencies (cause factor), since the latter might underestimate the burden of zinc deficiency. Evidence on the health burden of zinc is incomplete, leading to a significant reduction in reported health burdens in the Global Burden of Disease 2019. However, this reduction reflects merely a lack of data (Han et al., 2022; Hess et al., 2022), rather than a real-world reduction in zinc deficiency. Therefore, we adjust the GBD 2021 using an average DALY rate (per 100,000) and by adding other cause factors besides diarrheal diseases (Fischer Walker et al., 2009; GBD 2021 Risk Factors Collaborators, 2024; IHME, 2024b; Lassi et al., 2016; Liu et al., 2024; Veenemans et al., 2011).^2^

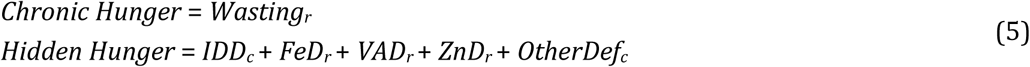

### 2.2 Relative nutrient intake increase

Crop breeding improves the nutrient supply by increasing yields and micronutrient contents of commodities, whereas fortification merely affects the micronutrient content, closing the nutrient intake gap in the diet. The extent to which fortification and biofortification can improve diets depends on (1) the crop improvement, (2) the importance of the respective crop in the diet, (3) the amount of food targeted and (4) the coverage of the food intervention. These factors collectively contribute to the increase in relative nutrient intake:

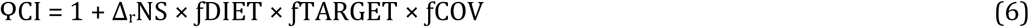

where (1) Δ_r_NS is the relative increase in nutrient supply provided by a specific commodity due to the food intervention, (2) ƒDIET is the proportion of total nutrient intake provided by a specific commodity, (3) ƒTARGET is the proportion of food consumption targeted (either the proportion of total nutrient intake provided by the production of a specific commodity for biofortification or the proportion of food industrially processed for fortification) and (4) ƒCOV is the proportion of the targeted food covered by the food intervention (either the proportion of the total area harvested that is planted by the new biofortified variety or the proportion of the industrially processed food that is fortified) (both depending on adoption).

The relative increase in nutrient supply, Δ_r_NS is as follows:

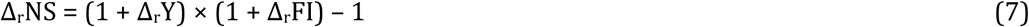

where Δ_r_Y is the relative increase in availability and affordability^3^ (%) (due to breeding for yield, agronomic improvements, food stamps, etc.) and Δ_r_FI is the relative increase in micronutrient content due to a food intervention (fortification or biofortification)^4^.

The proportion of total nutrient intake of calories and protein provided by a specific commodity, ƒDIET, can be obtained from FAOSTAT’s “Supply Utilization Accounts” (FAOSTAT, 2025). We correct refined sugar intake from FAOSTAT using Walton et al. (2023) and IHME (2024b) following GBD 2019 collaborators (2020).^5^ The intake of iron, zinc and vitamin A can be obtained by combining two commodity-level databases: (1) the food supply (in volumes) from FAOSTAT’s “Supply Utilization Accounts” (FAOSTAT, 2025) and (2) the Global nutrient conversion table (FAO, 2024). The food supply database is adjusted for outliers (using smoothing and averaging) and gaps. The food composition table is adjusted for outliers (using expert estimates) and fortification (using FFI, GAIN, IGN and the Micronutrient Forum (2025)).

The proportion of total nutrient intake provided by the production of a specific commodity, ƒPROD, can be calculated as the ratio of “production quantity” (in kg) from FAOSTAT’s “Crops and livestock products” over “food supply” (in kg) from FAOSTAT’s “Supply Utilization Accounts”. If needed, the ratio can be capped at 100%.

### 2.3 Relative nutrient intake gap

Zimmermann and Qaim (2004) hypothesised a rectangular hyperbolic relationship between health outcomes and nutrient intake (Figure 1A). Fischer et al. (2005) identified a strong empirical relationship between the prevalence of undernourishment and relative calorie intake (Figure 1B). We propose reversing this quadratic relationship to obtain estimates of the relative nutrient intake gap.^6^ Although Fischer et al. (2005) only identified the relationship for calorie intakes, the functional form can be applied to other nutrients after normalisation, assuming the same trend based on chronic hunger per capita versus calorie intake is the same for micronutrient deficiencies and intakes. Concretely:

1. Normalise the prevalence of undernourishment (from FAOSTAT) across all countries so that the maximum equals 1 (no limit to the minimum).
2. Obtain the relative calorie intake as the ratio of calorie supply over average dietary energy requirement (from FAOSTAT), and rescale so that the minimum (of the quadratic relationship) occurs at 1.

**Figure 1.**
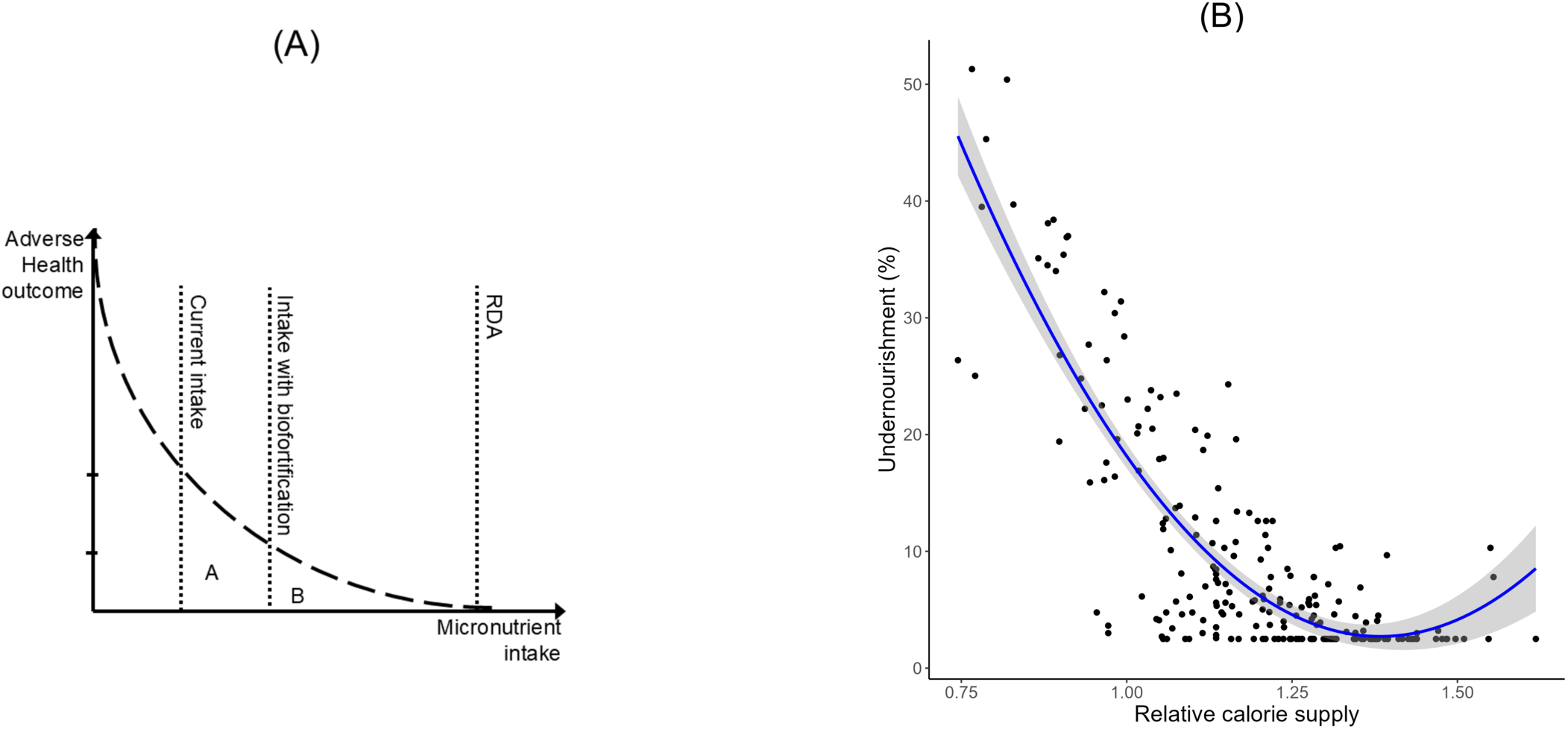
(A) Conceptual relationship between adverse health outcome and relative nutrient intake (source: Stein et al. (2005)). (B) Empirical relationship between the prevalence of undernourishment and relative calorie intake (source: own calculation using FAOSTAT following Fischer et al. (2005)).

The relative nutrient intake gap can then be modelled by (1) mathematically reversing the quadratic relationship between prevalence of undernourishment and relative calorie intake or (2) leveraging a regression model, enabling covariates.

#### 2.3.1 Quadratic model

The quadratic form can be inverted and used to calculate CI/RDA for given normalised levels of the burden of nutrient deficiencies per capita (from IHME):

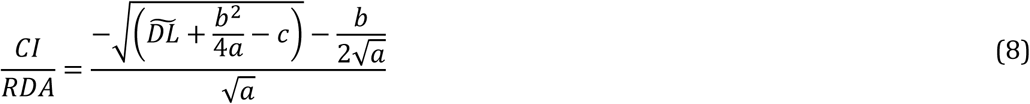

where 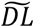 is the normalised disease levels (burden of nutrient deficiency), and a, b and c are the parameters of the quadratic relationship between the prevalence of undernourishment and the relative calorie intake (Figure 1B; Robinson et al., 2015, P. 28).

#### 2.3.2 Regression model (with controls)

Alternatively, we can regress relative calorie intake as a function of the prevalence of undernourishment and controls for food utilisation, like access to sanitation and intestinal infections.

#### 2.3.3 Comparison

Relative nutrient intake gaps are widely reported, but exhibit considerable variation across sources. For example, Lividini and Masters (2022) report that the apparent intake of vitamin A exceeds dietary reference values in Argentina, Mexico and West Africa. In contrast, IHME (2024b) data indicate that the burden of vitamin A deficiency in West Africa ranks among the fifth (highest) quintile globally, and Argentina and Mexico rank in the third (middle) quintile. Similarly, Lividini and Masters (2022) report that the apparent intake of bioavailable iron exceeds dietary reference values in Gabon and Papua New Guinea. In contrast, IHME (2024b) data indicate that the burden of iron deficiency in Gabon and Papua New Guinea ranks among the fifth (highest) and fourth (second-highest) quintile globally.

In this section, we compare three modelling approaches to quantify the relative nutrient intake gap: (1) a quadratic model, (2) a Random Forest (RF) model and (3) a Cubist model. The last two models use lack of access to sanitation (as a share of the population) (World Bank, 2025) and intestinal infections per capita (IHME, 2024b) as control variables. Overall, the cubist method reports the largest modelled intake gaps and the quadratic method the smallest. The random forest model gives less extreme estimates but is sensitive to generating outliers (for example, iron intake in Turkey and zinc and vitamin A intake in Nepal). Therefore, an ensemble model (with equal weights) across the three approaches gives the most balanced estimates.

Next, we compare the nutrient intake of iron, zinc and vitamin A using (1) our ensemble-modelled relative intake gap, (2) relative intake from Wang et al. (2023), (3) per capita intake from the Global Nutrient Database (GND) (GBD Nutrition Team, 2024; Schmidhuber et al., 2018), (4) relative intake and per capita intake from Passarelli et al. (2024), (5) per capita intake from the Global Dietary Database (GDD) (Global Dietary Database, 2018) and (6) relative intake^7^ and per capita intake from FAOSTAT (FAOSTAT, 2025). Figure 2 indicates that our modelled indicator has an overall high correlation with per capita GDP^8^, contrary to the intake metrics by Passarelli, Global Dietary Database and FAO, which show low to negative correlations with per capita GDP for at least one micronutrient. Correspondingly, many metrics—except the ensemble-modelled one—rank several low and lower-middle-income countries among the top 10 countries according to highest intake. Lastly, most relative intake metrics are not bounded between 0-100%, challenging their use in Equation 9. In sum, the ensemble-modelled nutrient intake gap shows the highest consistency with overall development, making it the most consistent and credible indicator of relative micronutrient intake globally.

**Figure 2.1A.**
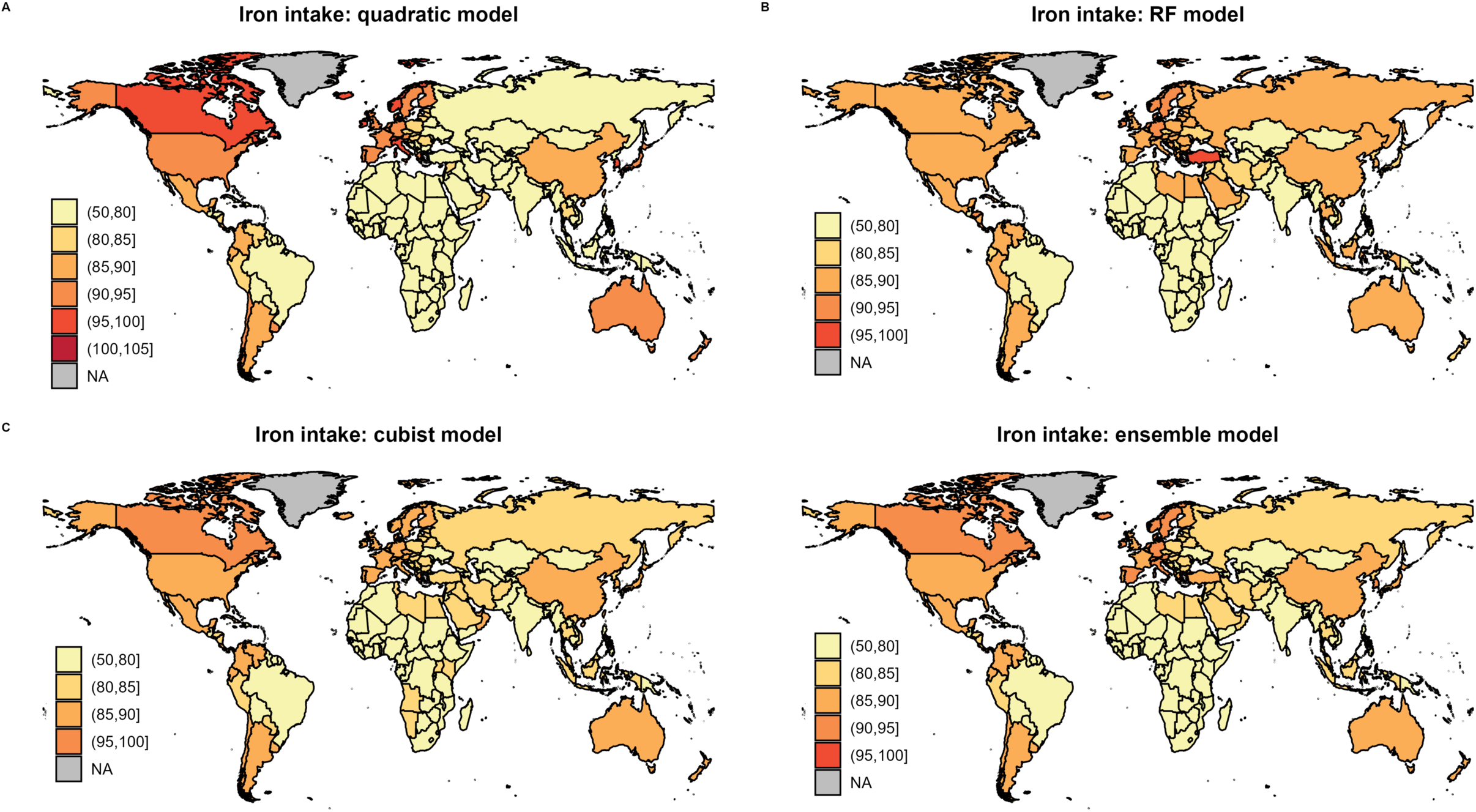
Modelled iron intake following different modelling approaches.

**Figure 2.1B.**
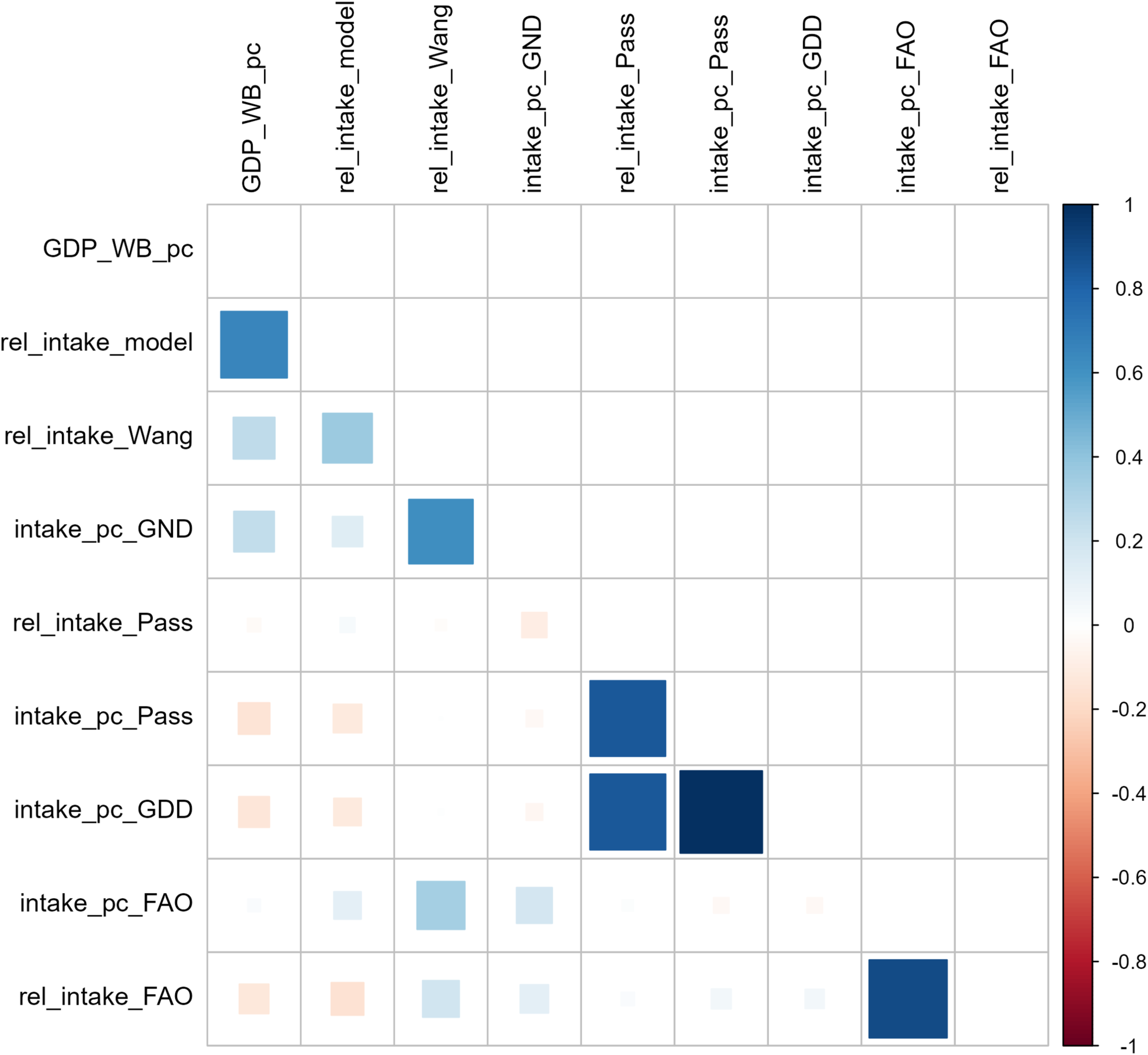
Correlogram of different iron intake metrics.

**Figure 2.2A.**
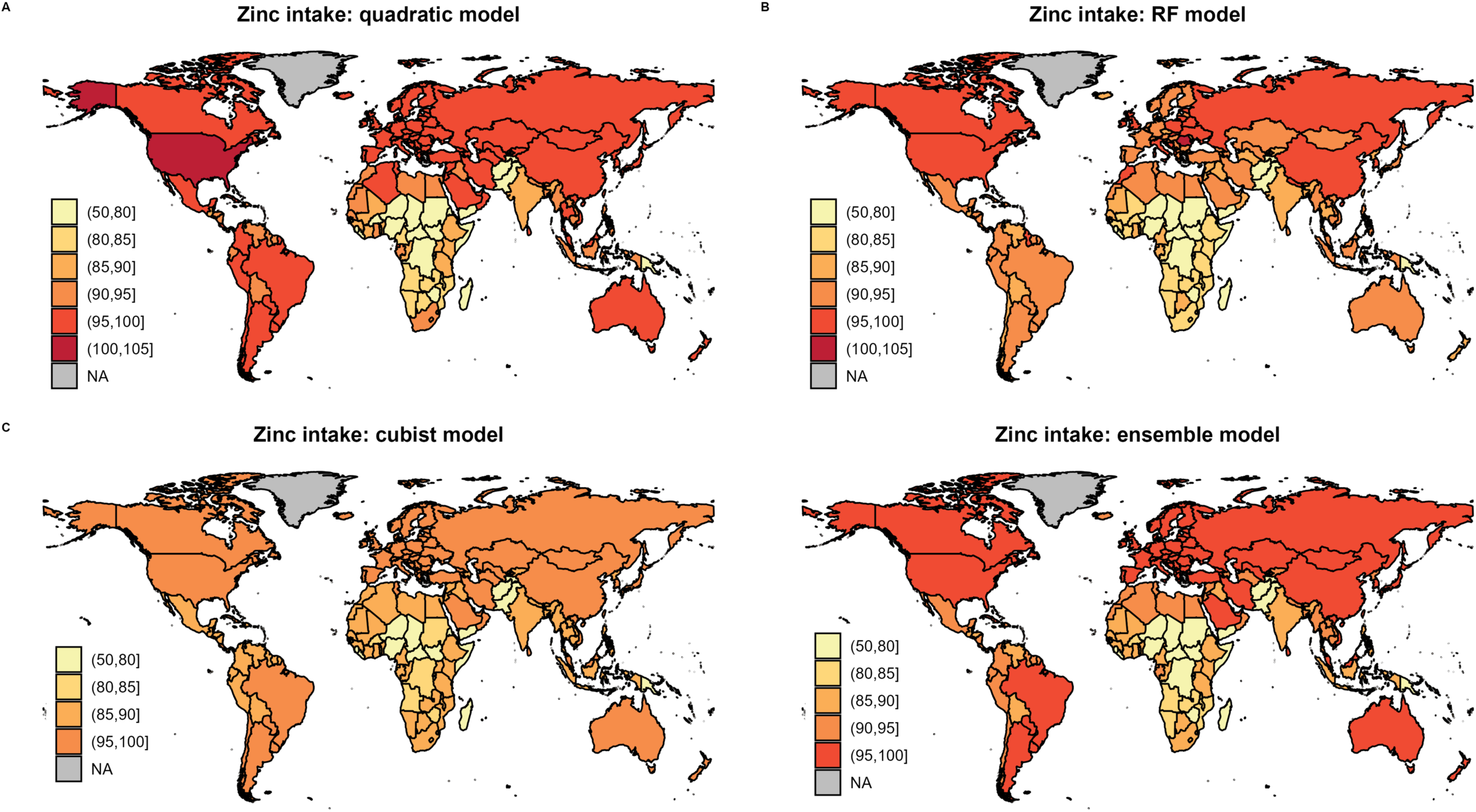
Modelled zinc intake following different modelling approaches.

**Figure 2.2B.**
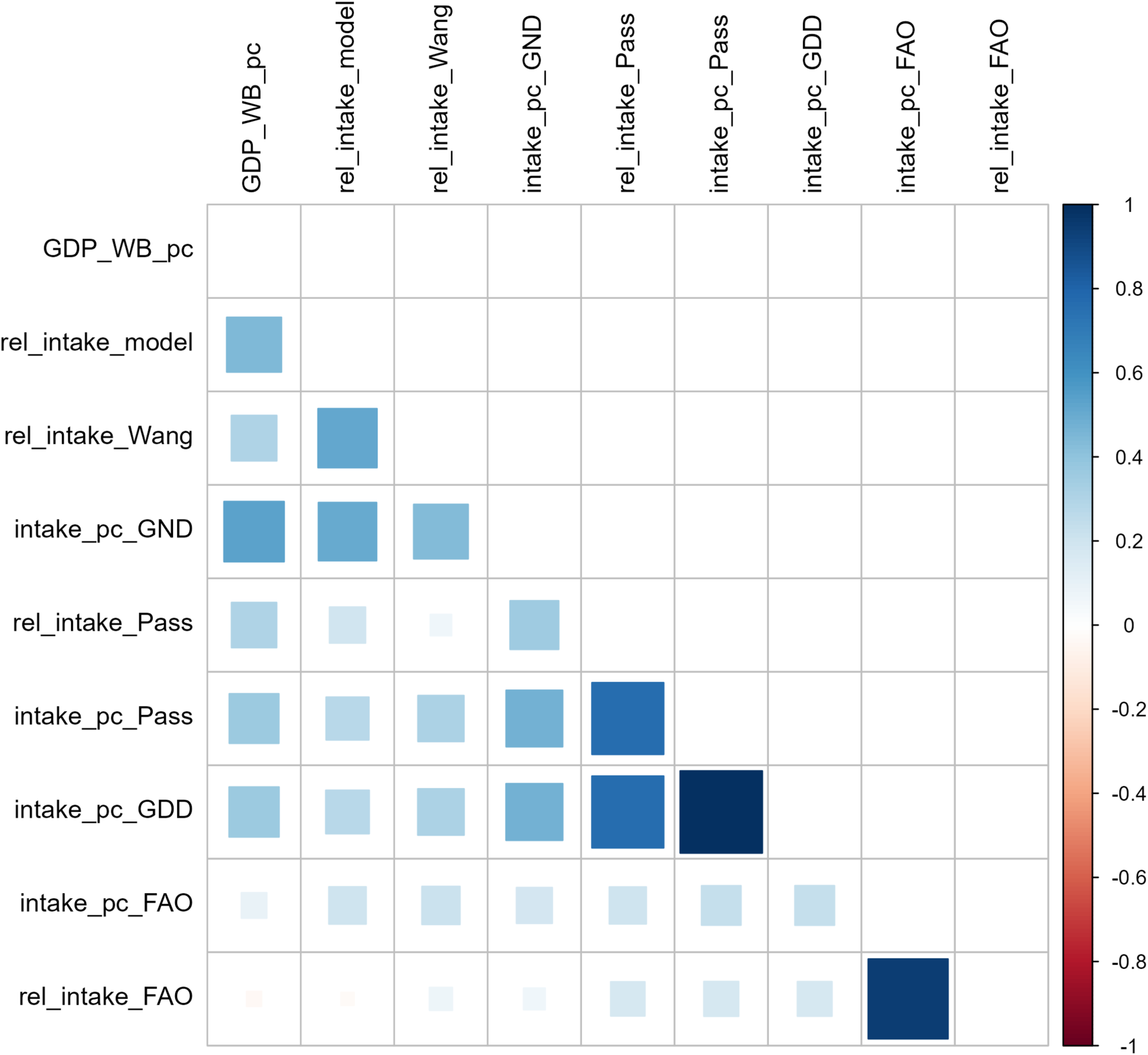
Correlogram of different zinc intake metrics.

**Figure 2.3A.**
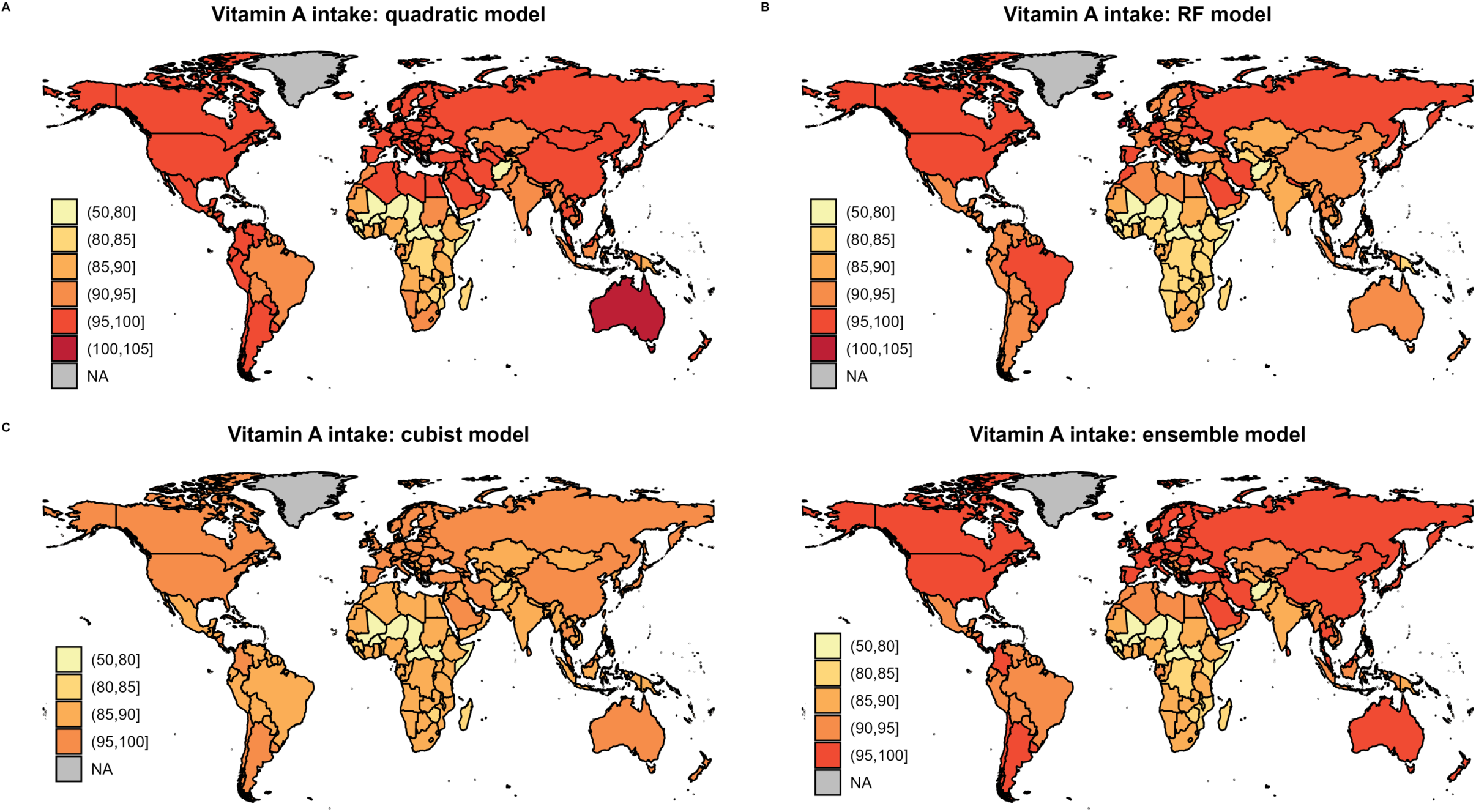
Modelled vitamin A intake following different modelling approaches.

**Figure 2.3B.**
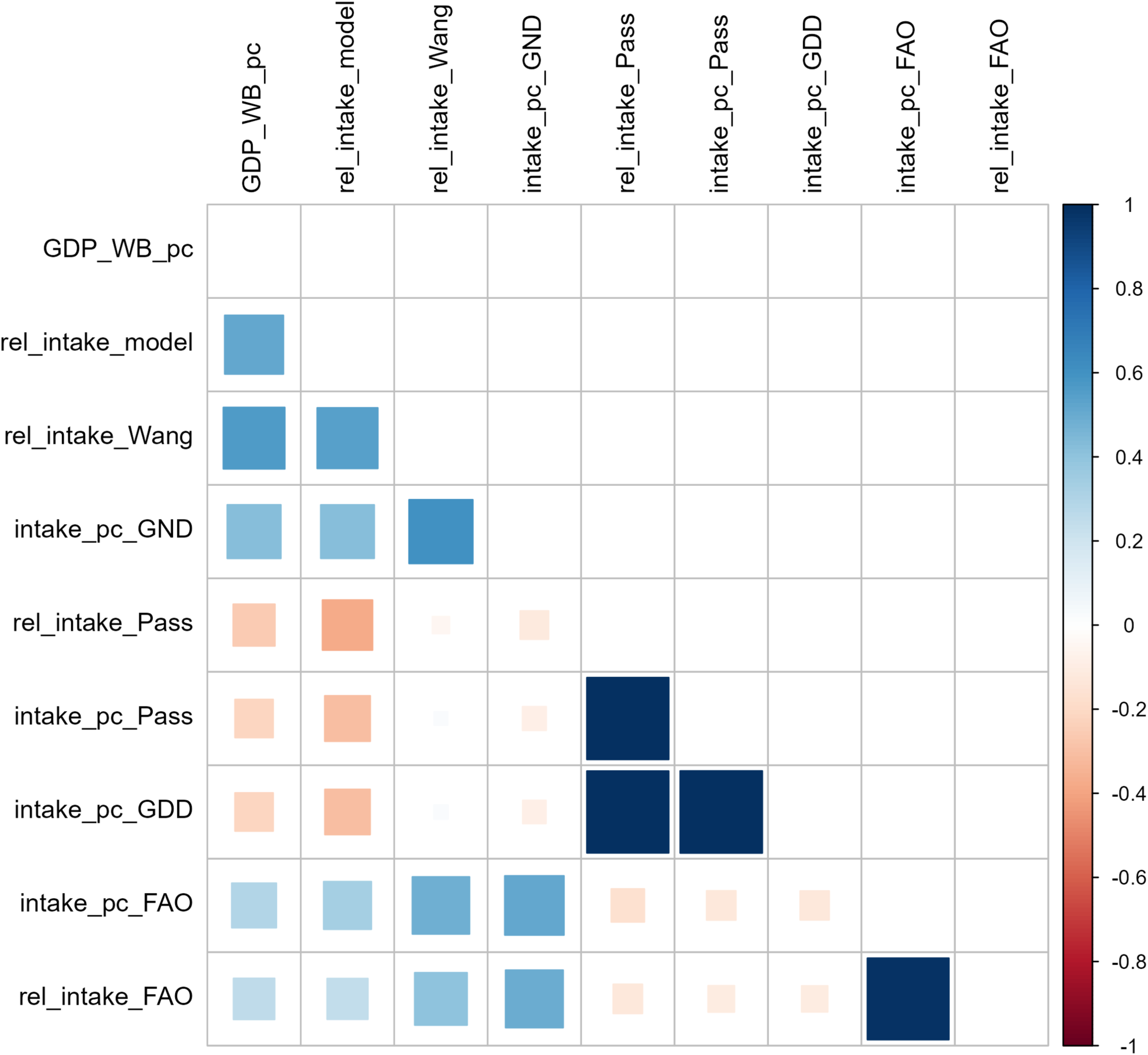
Correlogram of different vitamin A intake metrics.

### 2.4 Efficacy

We can now rewrite Equation 1 to incorporate the relative parameters introduced earlier:

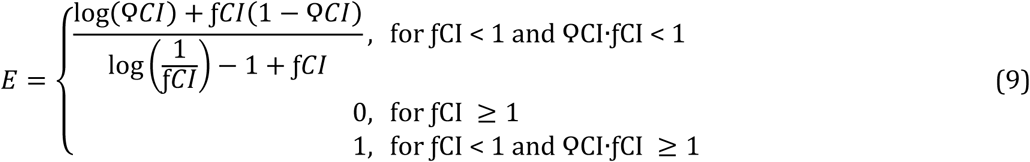

Equation 9 enables quantifying the health impacts of increases in nutrient supply, where ϘCI>1, ƒNS represents the relative increase in nutrient intake and E represents the relative decrease in the burden of hunger. However, adverse development trends like climate change, volatility in food prices, conflict and economic recessions might reduce the supply of nutrients available to consumers in lower-income countries. Therefore, we adapt equation 9 to incorporate decreases in yield:

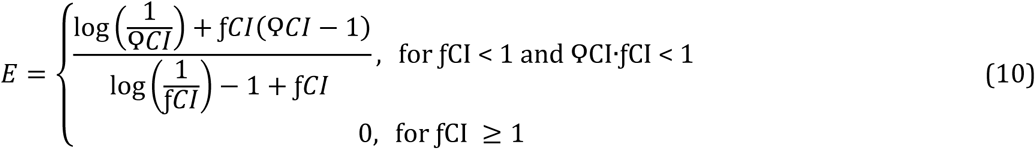

where ϘCI<1, ƒNS represents the relative decrease in nutrient intake and E represents the relative increase in the burden of hunger. All parameters used in the equations, except Δ_r_Y, are positive.

Considering the respective conditions, Equation 9 yields relative reductions in the health burden of hunger between 0 and 1. Figure 3 plots the efficacy rates versus the relative nutrient intake increase (subplot A) and versus the relative nutrient intake gap (subplot B). For high intake gaps, the efficacy rate increases almost linearly with the relative nutrient intake increase, whereas for smaller gaps, the efficacy converges slowly to 1, showing asymptotic behaviour. The efficacy rate drops to zero when the intake gap is closed. This reflects the higher scope of biofortification for reducing hunger in communities with considerable existing deficiencies.

**Figure 3.**
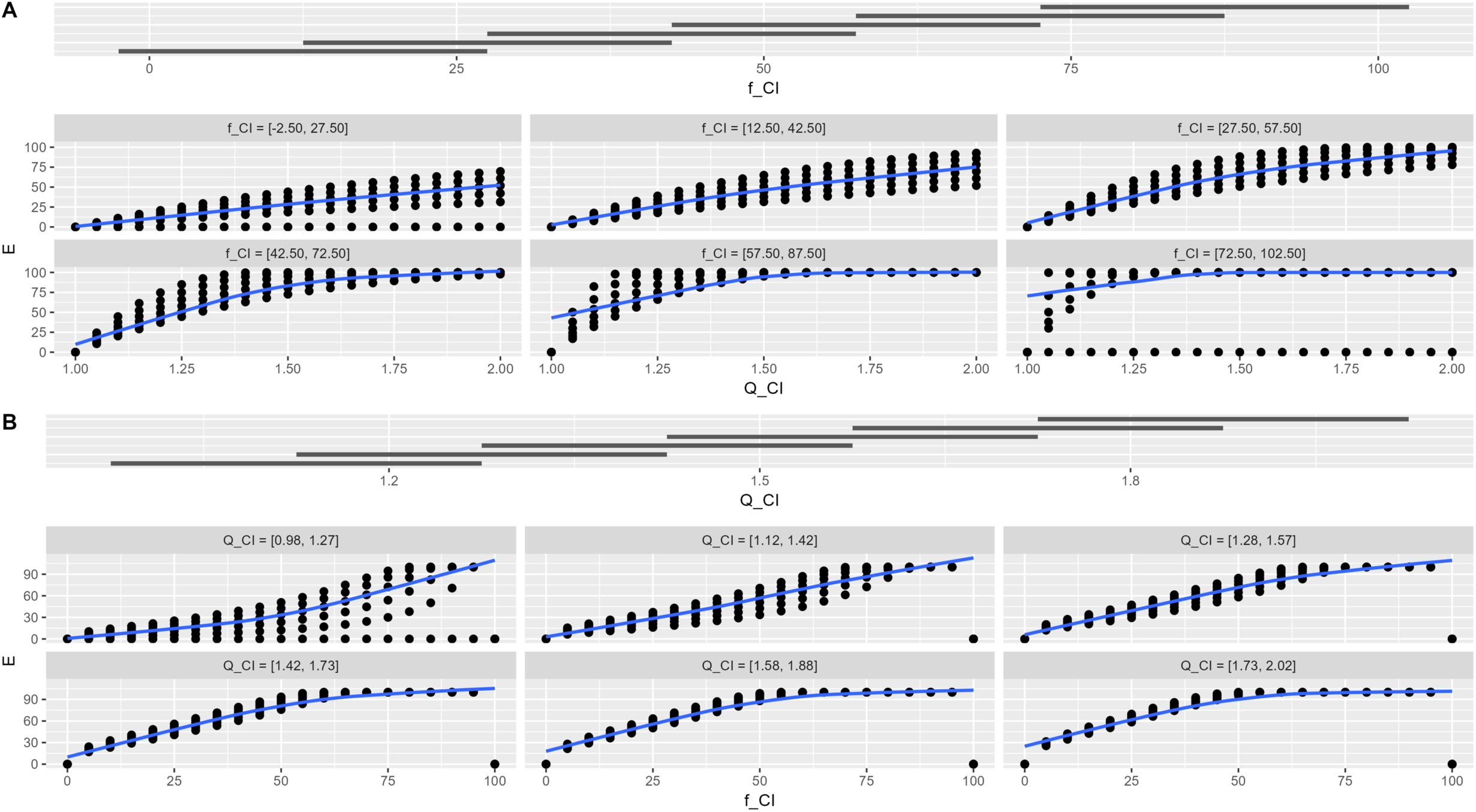
(A) Coplot of efficacy (E) versus the relative nutrient intake increase (Q_CI), controlling for the relative nutrient intake gap (f_CI). (B) Coplot of efficacy (E) versus the relative nutrient intake gap (f_CI), controlling for the relative nutrient intake increase (Q_CI).

Plot 2B ranges from 1 to 2, although most relative nutrient intake increases are not expected to exceed 1.125. The efficacy rate increases uniformly with the nutrient intake gaps, indicating it is easier to close smaller gaps, although the absolute burden of hunger avoided will be smaller. Like subplot A, subplot B evidences asymptotic behaviour and correlates zero efficacy rates with zero intake gaps. The asymptotic behaviour towards 1 reflects the conditions in Equation 9, without which the underlying formula might turn negative for CI > RDA or BI > RDA.

## Data Availability

All data used are available online from the sources listed in the paper.

## Acknowledgements

I sincerely thank my colleagues, Erick Boy and Victor Taleon (HarvestPlus), for their invaluable feedback and generous data sharing. Their support has greatly enhanced this research.

## Supplementary materials

**Table S1.**
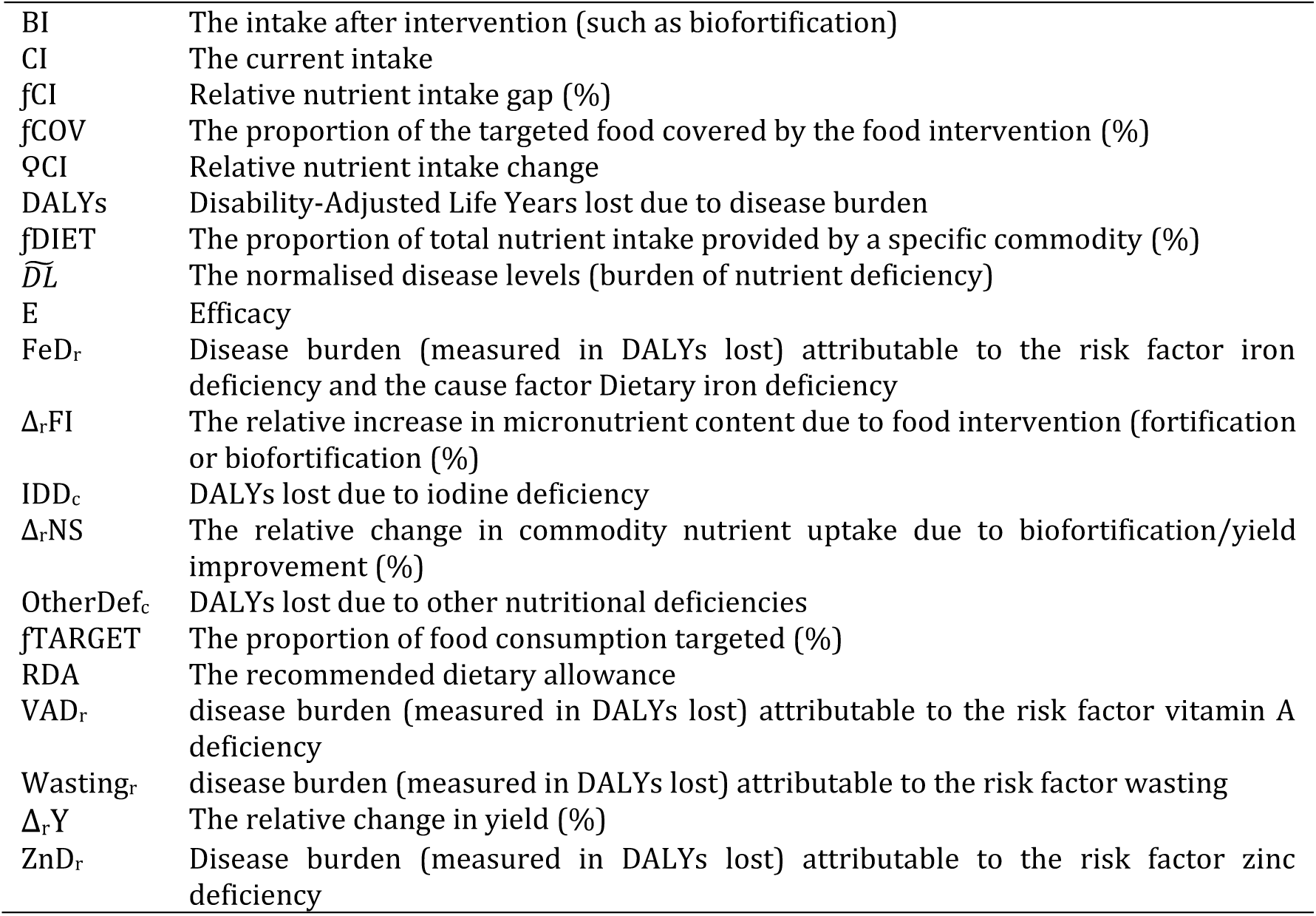
Index.

## Footnotes

2 This adjustment does not change the distribution of the health burden of zinc deficiency across countries, gender, age groups, etc. It merely corrects the ranking across nutrient deficiencies.

3 Assuming that increases in availability and affordability lead to increased consumption.

4 Bioavailability and retention rates might also be considered.

5 We use the reported sugar intake per person in the United States from Walton et al. (2023) and scale this to other countries using the normalised health burden of sugar-sweetened beverages intake from the GBD 2021. We then take the average between the refined sugar intake from FAOSTAT and the recalculated sugar intake.

6 Alternatively, if the distributions on intake and the Estimated Average Requirement (EAR) are known, the inverse of the prevalence of inadequacy (National Research Council, 1986) can be used.

7 Using average requirement estimates from the European Food Safety Authority.

8 For estimates of zinc and vitamin A deficiency, income per capita was used as a location-level covariate to inform supplementation estimates where data were absent (GBD 2021).

